# Epidemic Models with Random Infectious Period

**DOI:** 10.1101/2020.05.15.20103465

**Authors:** Germán Riaño

**Affiliations:** Amazon.com

**Keywords:** Epidemics, Stochastic, Transient Little Law, Random Infectious Period

## Abstract

In this paper, we present an extension to the classical SIR epidemic transmission model that uses any general probability distribution for the length of the infectious period. The classical SIR model implicitly requires an exponential distribution for the length of this period of time. We will show how a general distribution can be easily taken into account using the Transient Little Law and present numerical methods to solve the model in an efficient way. Our numerical experiments show that in the presence of a more realistic distribution, with lower variability than the exponential distribution, the size of peak of infected individuals on the graph will be higher and occur earlier. Conversely, a higher-variability distribution will lead to a lower peak that takes longer to dissipate. We also discuss some extensions to the basic model, to include variants like SEIRD and SIS. These findings should have profound and important consequences in the design of public policy.

## 1 Introduction

The advent of the COVID-19 pandemia has placed many challenges on society. One of them is deciding the proper public policy in the face of many diverse predictions. Predicting the size, and ultimate impact of the pandemia, is not easy, because there are still many unknowns. We think, however, that there is a contributing factor that, to the best of our knowledge, has not been identified in academic literature: most models used for prediction are some variant of the SIR model, that does not explicitly consider the probability distribution for the length of the infectious period, but rather assume a recovery rate proportional to the number of infectious individuals. As we will see, this is akin to assuming an exponential distribution.

The purpose of this paper is not to give a prediction of COVID-19 evolution for any particular country or region, but rather present extensions to classical mathematical models of epidemic transmission using a general distribution for the length of the infectious period. We will show that the right choice of distribution does have an important impact on the predictions made by the model.

As a motivation for the modeling techniques presented in this paper, consider the classical SIR model [1, 2, 3]. The population is split in three groups: the Susceptible (S) are all individuals in a population that have not been infected; Infectious (I) are those individuals that have acquired the disease and are assumed to be infectious from the moment they get it; finally, the Removed (R) are the individuals that have either recovered or died, and are assumed to have gained immunity and, therefore, will not be infected again. It is assumed that the total population remains constant, so there are no births and no deaths for other causes other than the epidemic itself. The dynamics of the epidemic are described by the following set of Ordinary Differential Equations (ODE)

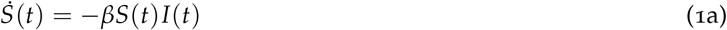

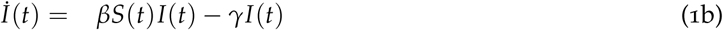

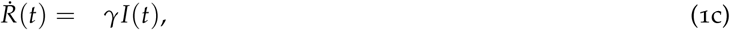

subject to initial conditions *I*(0) = *I*_0_, *S*(0) = *S*_0_, and *R*(0) = *R*_0_. The dot denotes the derivative with respect to time. In this paper, we will adopt the convention that the previous quantities are measured as a percentage of the total population, so *S*_0_ + *I*_0_ + *R*_0_ = *S*(*t*) + *I*(*t*) + *R*(*t*) = 1. The term *βS*(*t*)*I*(*t*), in equations (1a) and (1b), tells us that the rate of incidence of new infections is both proportional to the number of infected individuals, *I*(*t*), and also to the proportion of available individuals, *S*(*t*), and this makes sense. The term *γI*(*t*), however, is problematic. It implies that in the next *∆t* any of the infected individuals will recover with the same probability *γ∆t*, regardless of how long ago they acquired the disease. A random variable that exhibits this behavior is said to have a *memory-less property*. This, in turn, implies that the length of the disease must follow an exponential distribution with mean *γ*^−1^, since it is well-known that the exponential is the only distribution that follows this property (see, e.g., [4]).

We will show how to incorporate in the model an arbitrary distribution for infectious period. The resulting model is not a set of ODE, but it is still easy to compute in a few seconds. To the best of our knowledge, our results are new. The reader can see other techniques that have been applied to stochastic modeling of epidemics in the surveys [5] and [6], and references therein.

The remainder of the paper is organized as follows: in Section 2 we present the SIR-G model, a SIR model that incorporates any general distribution for the infectious period; in Section 3 and 4 we present an algorithm to efficiently solve the model and discuss numerical experiment with diverse distributions; in Section 5 we present results using Phase-Type distributions, and generalize it in Section 6 to a setting with multiple stages with arbitrary duration (a SEIRD model is discussed as a particular instance of such a model); in Section 7 we discuss the limit behavior of the models presented and prove that the number of individuals eventually infected will be the same as in the classical SIR model; finally, we discuss how the modeling techniques can be extended to other models and give concluding remarks.

## 2 SIR-G: A SIR Model with General Random Infectious Period

### 2.1 The Transient Little Law

In this section we will briefly discuss the Transient Little Law (TLL)[7], since it will be the main building block for the models discussed later. Consider a general system to which entities arrive, spend some time and eventually leave. In our case, the *system* will be the collection of infectious individuals in a population, but, in general, the system can be a bank, a call center, etc. Let *{N*(*t*), *t ≥* 0} represent an stochastic process that counts the cumulative number of arrivals up to time *t*, and will denote its expected value as *Λ*(*t*) = *E*[*N*(*t*)], and the corresponding derivative as *λ*(*t*). Arrivals spend a time *T* characterized by the distribution *G*(*t*) *≡ P*{*T* ≤ *t*}. For convenience, throughout this paper, we will often use the survival function, 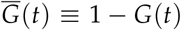. For our case, we will assume this time is independent of the arrival process, although those assumptions are not required for the TLL. For the moment, assume at time *t* = 0 the system is empty. At any later time, *t* > 0, the number of expected arrivals that have occurred so far, *Λ*(*t*), will be split as *Λ*(*t*) = *I*(*t*) + *R*(*t*), where *I*(*t*), is the expected value of entities still in the system, and *R*(*t*) is the expected value of departures. The TLL tells us that

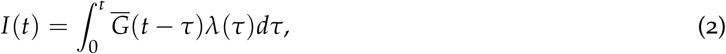

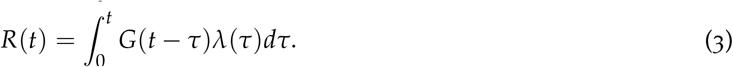

See the proof in [7]. The result is very general and does not require any type of assumption with respect to the arrival process. Also, we do not need to know any specifics about the stochastic process that characterizes the number of entities inside the system, since the TLL is a relationship for the *expected* values.

The previous equations ignore the individuals already in the system at time 0, so we will now show how to take them into account. Assume that we know the initial values of *I*_0_ = *I*(0) and *R*_0_ = *R*(0). However, we will usually not know how long they have been in the system. It will be natural to assume that, for a particular individual already infected at time 0, instant *t* = 0 is a random incidence during the length of *T*. From Renewal Theory results, we know that the remaining time will follow the so-called *equilibrium residual distribution* (see, e.g., [4]), given by

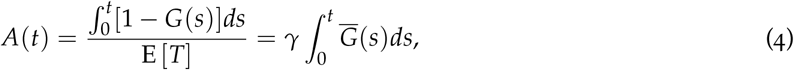

where *γ*^−1^ = *E*(*T*), i.e., *γ* would be the parameter of the corresponding exponential with the same mean. A random individual among the initial *I*_0_ would have already left the system at time *t* if this residual time is less than *t*, which occurs with probability *A*(*t*) and would still be inside with probability 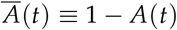. Therefore, to consider the initial conditions, we modify (2) and (3), to get

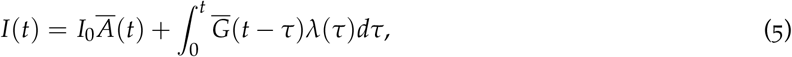

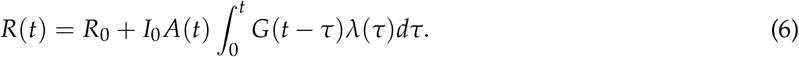

Notice that we can verify *I*(*t*) + *R*(*t*) = *I*_0_ + *R*_0_ + *Λ*(*t*), as expected.

### 2.2 Integral SIR-G Model

In this section, we will present a model that generalizes the classical SIR model (1a)-(1c) in a way that allows the use of a general distributions for the infectious period, which may or may not be an exponential. We will apply the Transient Little Law, i.e., ((5)) and ((6), to this system. For our case, the so called *system* will be the pool of infectious individuals, and, therefore, *I*(*t*) is the expected value of infectious, and *R*(*t*) the expected value of removed individuals, either by death or recovery. As in the classical SIR model, we will assume that the rate of change of *Λ*(*t*) is given by

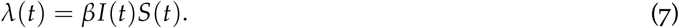

Therefore, (5) and (6) become

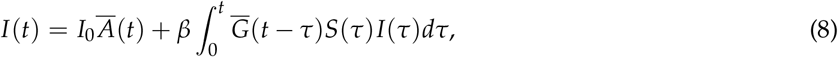

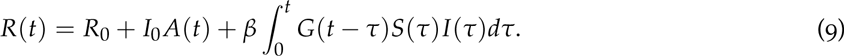

Equations (8) and (9) can be used in conjunction with the integral of (1a) to fully characterize the behavior of the epidemic. Therefore, we have to solve the following system of equations

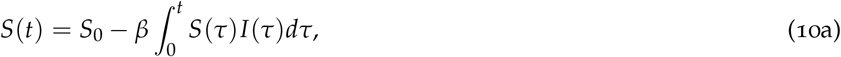

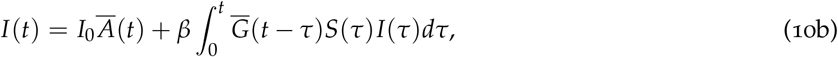

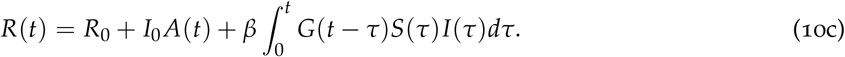

Note that *S*(*t*) + *I*(*t*) + *R*(*t*) = *S*_0_ + *I*_0_ + *R*_0_ = 1, as expected. We will call this set of equations the **Integral SIR-G Model**.

### 2.3 Integro-differential SIR-G Model

The SIR-G model can be also presented in integro-differential form, and that is the way we will use for computation in Sections 3 and 4, by taking derivatives with respect to time of Equations (10a)-(10c). The derivative of the integral in (10b) can be computed using Leibniz rule, and using 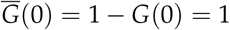, as follows

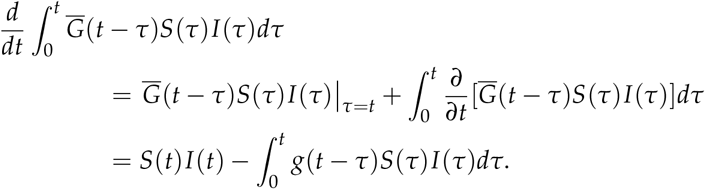

Therefore, taking derivatives in equations (10a)-(10c), we get the SIR-G in integro-differential form:

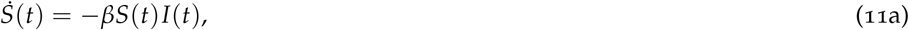

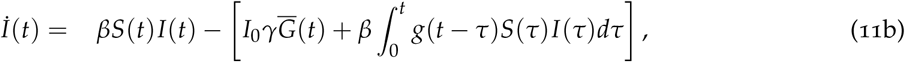

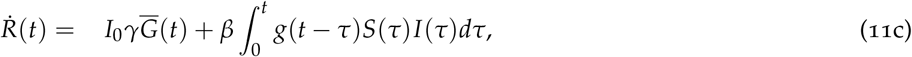

subject to *S*(0) = *S*_0_, *I*(0) = *I*_0_ and *R*(0) = *R*_0_. Here we have used 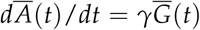, which can be seen from (4).

### 2.4 Recovering the Classical SIR with Exponential Distributions

We finish this section proving that the previous set of equations is equivalent to the classical SIR model (1a)-(1c) when the distribution *G* is exponential. For the exponential case *g*(*t*) = *γe*^−*γt*^ and 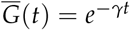, so 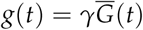. Also, plugging 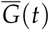 in the residual distribution (4), we can see 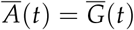. In other words, the residual time is also exponential, which should be expected because of the *memory-less* property. Now, the terms in brackets in (11b) can be computed as

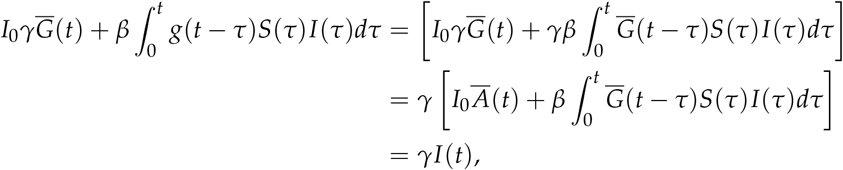

where the last equation comes from (10b). Plugging this into (11b) and (11c), we recover the original SIR model ((1a)-(1c)), showing, once again, that the classical SIR model is valid only when the distribution is exponential.

### 2.5 Computing Deaths and Recoveries Independently

In the previous model, *R*(*t*) counts all individuals that were removed from the pool of infectious individuals. This contains both the ones that died and those that recovered from the disease. If the time of the infection is the same for both of these groups, then the number of deaths and recoveries can be computed simply as *R_d_*(*t*) = *p_d_R*(*t*) and *R_r_*(*t*) = *p_r_R*(*t*), where *p_d_* and *p_r_* are, respectively, the death rate and recovery probabilities (and, of course, *p_d_* + *p_r_* = 1). However, we can also have two different distributions for people that die and for those that recover, say *G_d_*(*·*) and *G_r_*(*·*), and then, in this case, *G*(*·*) will be the mixture

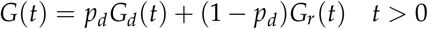

Each of these distributions will have its corresponding residual distributions, say 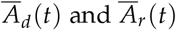. We can also modify (10c) to compute the cumulative number of deaths and recoveries, independently, as

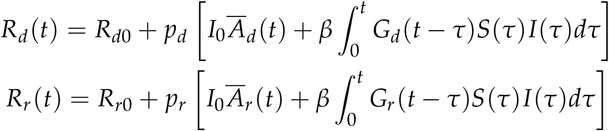

Notice that *R_d_*(*t*) + *R_r_*(*t*) = *R*(*t*), as expected. Similar manipulation can be done to obtain differential versions.

## 3 Computation Algorithms

In this section, we discuss algorithms to compute the SIR-G model. We will focus on the integrodifferential version, in particular using equations (11a) and (11b). The equation for *R*(*t*) can be ignored, since it can be computed later as *R*(*t*) = 1 *− I*(*t*) *− S*(*t*). In other words, we focus only on determining functions *S*(*t*) and *I*(*t*).

## 3.1 Simple Forward Computation

In the most simple algorithm, we sub-divide the timeline in intervals of size *δ*, and compute *S* and *I* only at those points, i.e., *S_k_* = *S*(*δ_k_*) and *I_n_* = *I*(*δ_k_*). Discretizing (11a) and (11b), we get the computation scheme in Algorithm 1.

### Algorithm 1

SIR-G algorithm

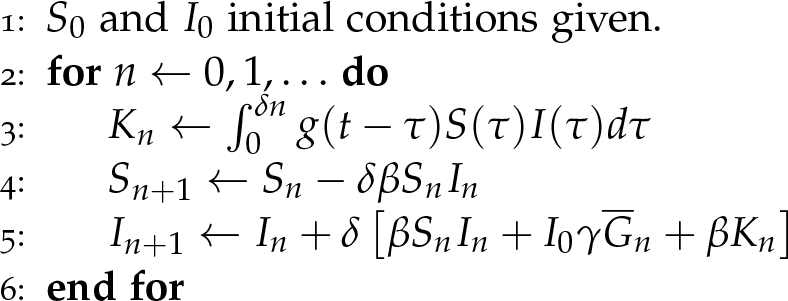

Of course, the difficult part is computing the integral. Notice that this integral depends only on parts of *I*(*t*) and *S*(*t*) that already have been computed, but the function is only known at discrete points. One could use a computation scheme like Simpson rule, readily available in many libraries like SciPy [8]. However, for some distributions, *g*(*t*) can be very high when *t* is close to 0, and that would pose a challenge for the precision and stability of the integral computation. Therefore, we devised a robust algorithm to compute the integral, converting *I*(*t*)*S*(*t*) into a piece-wise linear function and then computing the exact integral. Let *B*(*t*) ≡ *S*(*t*)*I*(*t*). Notice that the integral can be written as

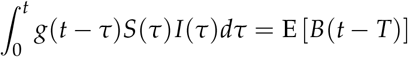

Therefore, we can frame this problem as finding the expected value of a piece-wise linear function *f* (*x*) = *B*(*t − x*) (keep in mind that *t* is fixed). See the details in Appendix A. Our implementation is available at GitHub[9].

## 4 Numerical Experiments

### 4.1 Analysis with Covid-like data

In this section, we study the behavior of the SIR-G system using diverse distributions. First, given the current prominence of the spread of Covid-19, we run the model with parameters close to those that have been reported for this disease. In particular, we will use the parameters reported in [10]: we assume that *T* is distributed as a *Normal*(*mean* = 4.5, *SD* = 1), and the basic reproductive number, *ℛ*_0_ = 2.2. *ℛ*_0_ represents the number of individuals infected directly by an individual that is infected. In the classical SIR model, *ℛ*_0_ = *β*/*γ*. Our purpose here is not to present a fit for Covid-19 dynamics in any particular geography, since that is outside of the scope of this paper. Rather, if we assume that the given parameters are valid for a particular hypothetical epidemic, that we will call DIVOC, we will show how different the dynamics will be for the SIR-G model compared to the classical SIR model. In Figure 1, you can see how these models behave. The model that uses a realistic distribution (dotted lines) has a peak higher and earlier. The classical SIR model predicts size of the peak to reach 19% of the population and occur on day 50. However, in the SIR-G model we see that the peak is reached much earlier on day 36 and will reach 36%. This would have important consequences on the strain placed on medical facilities and the number of ICU’s beds and respirators needed. Interestingly enough, the limit value of *R*(*t*) is the same in both cases. Therefore, in both models the number of people that ever gets infected is the same (and consequently, assuming all patients can get the same quality of care, the number of deaths will be identical as well). In Section 7, we provide a mathematical proof that this result will hold in general, regardless of the distribution.

**Figure 1:**
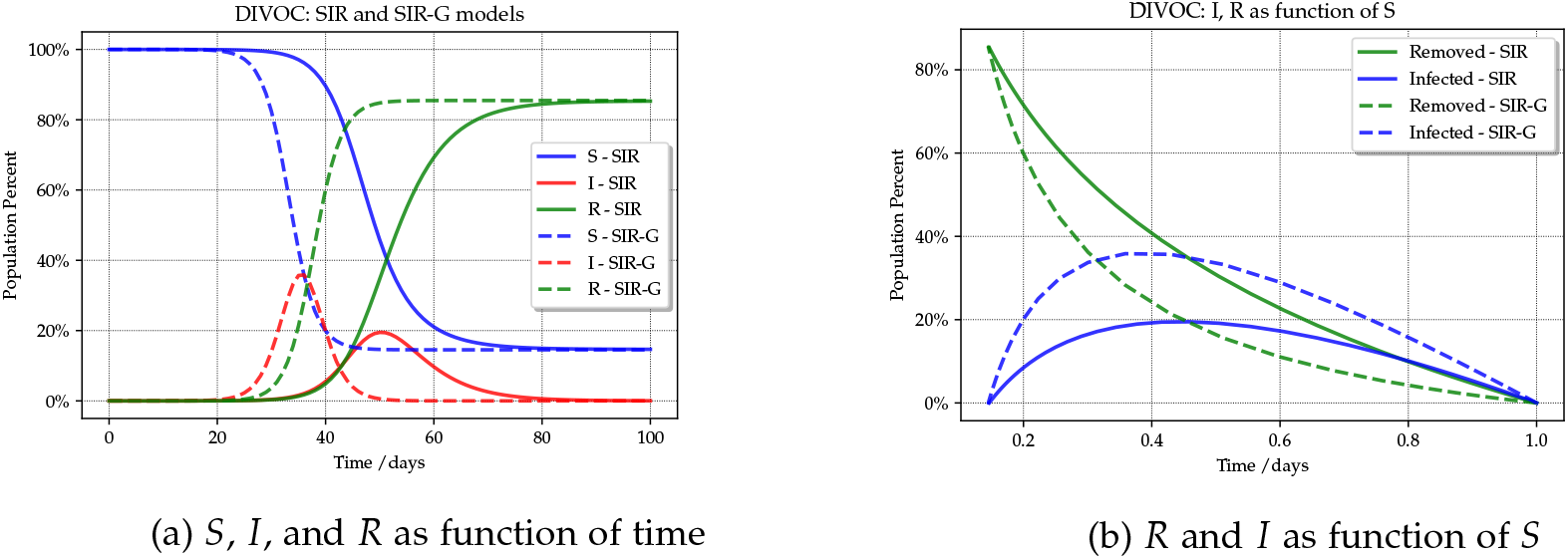
Comparison of classical SIR with SIR-G

Another way to see this result is as follows: if the distribution of the infectious time is correctly estimated based on the evolution of individual observed cases, but *ℛ*_0_ is based on fitting a classical SIR model to the number of observed cases (or deaths), then the analyst would predict a much higher *ℛ*_0_ than the real number. In other words, if the analyst sees the dotted line he or she would conclude that the disease spreads faster than what it actually does. Additionally, the predicted number of deaths will be higher than the real figure.

### 4.2 Impact of Variability in SIR-G

In this section, we will use diverse Gamma and Log-normal distributions. For the experiment, we always used distributions with the same expected value, but changed the variance. We fixed the coefficient of variability, *CV* at values 0.5, 1 and 2. (*CV* is defined as 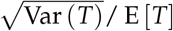; notice that for the exponential *CV* = 1). In Figure 2, we can see that a higher variability causes a lower and later peak as compared to the classical SIR model. Conversely, if variability is lower, the peak comes higher and earlier. The highest peak is obtained with a constant time for the infectious period (solid blue line in the graph). We also see that for higher variability distributions, the CV does not tell us the full picture, since the Gamma and Lognormal with the same CV produce different results. In other words, higher moments will have an effect on the evolution of the system.

**Figure 2:**
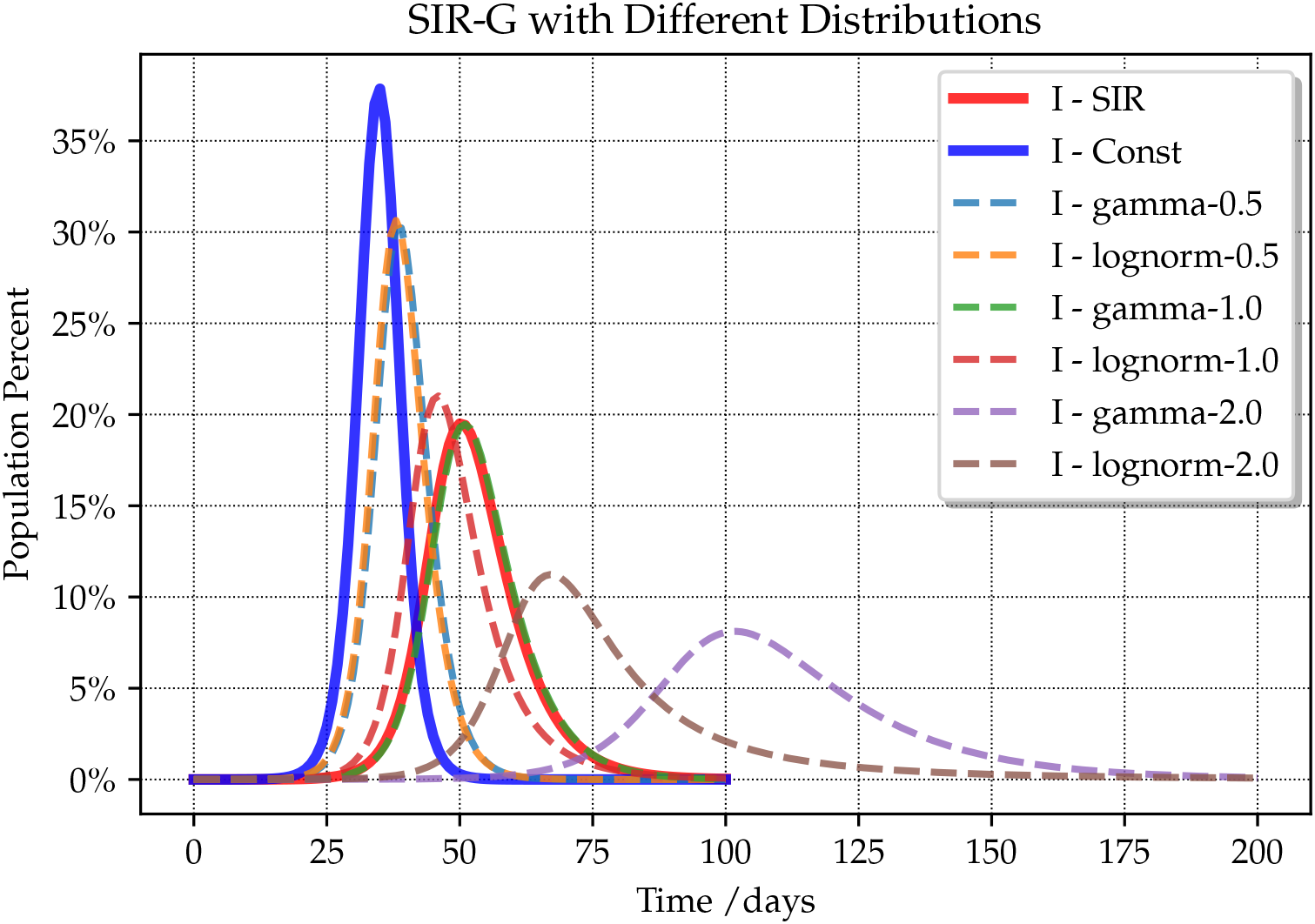
SIR-G for different variability levels and diverse distributions.

## 5 Epidemic Model with Phase-Type Distributions

In this section, we will describe a SIR-G model for the particular case when *T* has a Phase-Type distribution (PH) [11, 12]. Our interest in PH distributions is two-fold. First, it can be shown that the family of PH distributions is dense, in the sense that any positive random variable can be approximated by a PH distribution [11]. Also, the family of PH distributions is closed under many operations. In particular, as we will see, the residual equilibrium distribution in (4) is a PH distribution. There are well established methods to fit data or moments to create a PH distribution (see e.g. [13, 14, 15]), and there are libraries that allow users to easily fit data to distributions and manipulate the variables with relatively little coding ([16, 17].)

A PH variable can be thought of as the absorbing time in a Continuous Time Markov Chain (CTMC) with a single absorbing state. To build a PH distribution, imagine a CTMC with states *{*0, 1, …, *n*}, where the state 0 is the only absorbing state. The CTMC behavior is characterized by generator matrix and initial condition vector given by

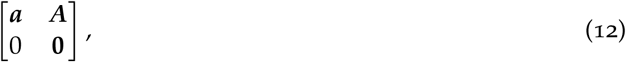

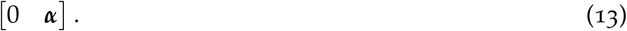

Here, ***A*** is an *n* × *n* matrix that corresponds to the transient part of a CTMC and ***α*** is the row vector of initial conditions. If the variable is purely continuous, these probabilities must add up to 1, so ***α***1 = 1, where **1** is a *column* vector of ones. The behavior of a PH can be thought of this way: a Markov chain chooses one initial state *i* (or phase) with probabilities *α_i_*, then moves among phases according to rates *A_ij_*, and, eventually, it’s absorbed, with rate *a_j_* from some state *j*. The phases in the PH distribution do not necessarily have a physical interpretation: they can come as a consequence of the data fitting process.

Since the matrix in (12) is a generator for a CTMC, the rows must add up to zero, and hence

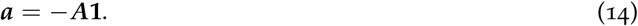

It can be shown that the CDF for the time until absorption, i.e., the PH distribution, is given by

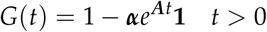

From this, we can derive the PDF, expected value, and equilibrium distribution as

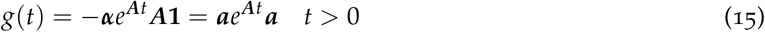

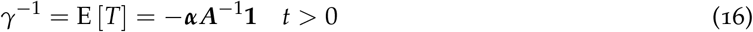

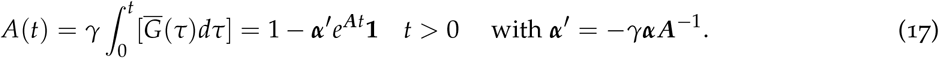

Notice that the residual equilibrium distribution is also a PH with the same matrix but different initial conditions ***α***^′^(it can be shown that ***α***^′^ ≥ 0 and ***α***^′^**1** = 1.). Typical examples of PH distributions include the Exponential, Erlang and Hyper-exponential. In Table 1, we show the corresponding representation.

**Table 1:**
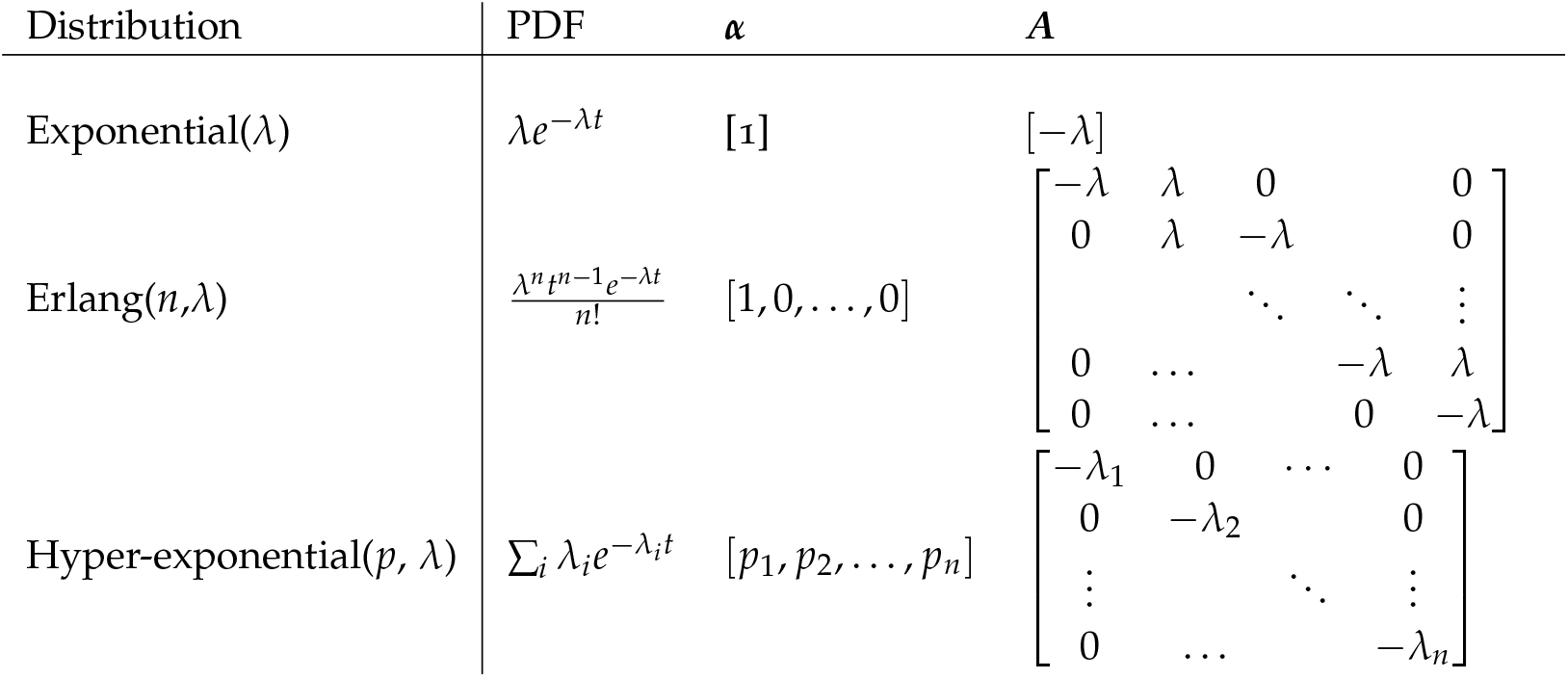
Some common examples of PH distributions.

Plugging these results in (8), we get

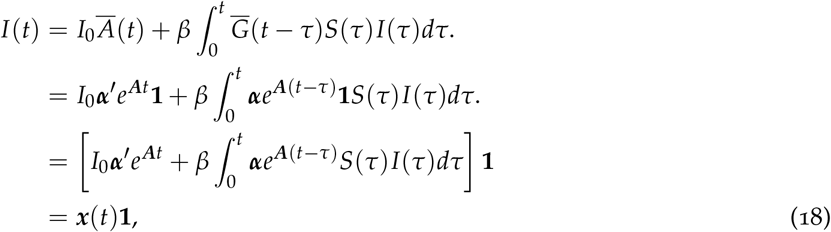

where ***x***(*t*) is the equation inside the braces; it is a *row* vector whose *i*-th component represents the fraction of infected individuals currently in phase *i*, and *I*(*t*) = ***x***(*t*)**1** is just the sum. We can take the derivative of ***x***(*t*), using Leibniz rule, to get

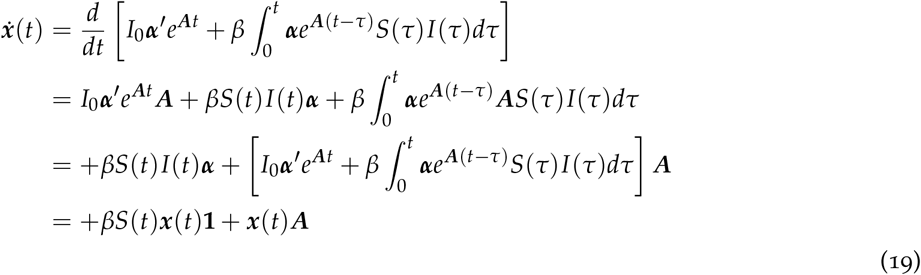

With similar manipulation, it’s easy to show that *Ṙ*(*t*) = *−**x***(*t*)***A*** = ***x***(*t*)***a***. Therefore, for the case when *G* is a PH distribution, we can build a new model that we will call SIR-PH. The equivalent of equations (10a)-(10c) becomes

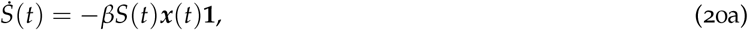

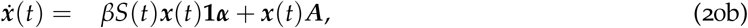

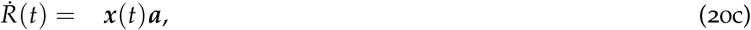

subject to initial conditions *S*(0) = *S*_0_, ***x***(0) = *I*_0_***α***^′^ and *R*(0) = *R*_0_. This model will have more equations than the original SIR-G, but it can potentially be faster to solve since it is an ODE and there will be no need to compute integrals. Notice the resemblance with the original SIR in equations (1b)-(1c). Of course, the classical SIR will be a particular case when ***a*** = *−**A*** = *γ*. Another advantage of this representation is that it allows us to study limit behavior of the system, but we will delay that until after the next section.

## 6 A Multi-stage Epidemic Model

In this section, we generalize the SIP-PH model. Assume that an infected individual goes through different stages, like in the previous model, and once he or she is infected it would evolve according to a CTMC characterized by ***A***, until eventually is removed from stage *i* with rate *a_i_*. However, we will assume that there is an infection rate, *β_i_*, that might be different for the different stages (for example, there might be an incubation stage where the individual has been exposed, but is not yet contagious). The number of new infectious generated by stage *i* is given by

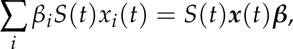

where ***β*** is a *column* vector with the different rates. These new infectious individuals start on stage *j* with probability *α_j_*, as before, so the rate of arrival of new infected to stage *j* is given by the *j*-th component of vector *S*(*t*)***x***(*t*)***βα***. We now replace this infectious dynamics in (20a)-(20c), to get the following model that we will call SIR-PHG

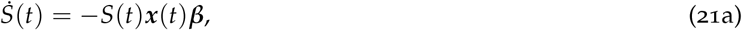

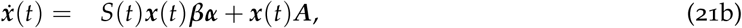

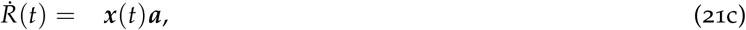

This model can be generalized even further by replacing ***βα*** by a matrix ***B***, such that *B_ij_* represents the rate of contagion by individuals type *i* that cause an infection that starts in *j*.

### 6.1 SEIRD-PH Model

The fact that the evolution of the disease uses a CTMC might seem overly restrictive, since it requires the stages to be exponential. However, that is not the case. With the help of PH distributions we can combine multiple phases to make different stages whose sojourn times are PH random variables. As an illustration, consider the following *SEIRD* model. Susceptible individuals are infected at rate *β* at which point they are considered *exposed* but are not yet infectious (see Figure 3). They remain in the exposed stage during a time *T_e_* that has distribution *PH*(***α**_e_, **A**_e_*) after which they will be infectious. With probability *p_d_* the individuals would die after a time *T_d_* that is *PH*(***α**_d_, **A**_d_*), and with probability *p_r_* = *a − p_d_* they will recover, after a time *T_r_* that is *PH*(***α**_r_, **A**_r_*). This model can easily be placed in the formalism of equations (21a)-(21c). The total time will be either *T_e_* + *T_r_* if the person recovers, or *T_e_* + *T_d_* if the person dies. Using the operations for mixtures and sum of PH random variables [12], the matrix ***A*** and initial conditions ***α*** will be given by

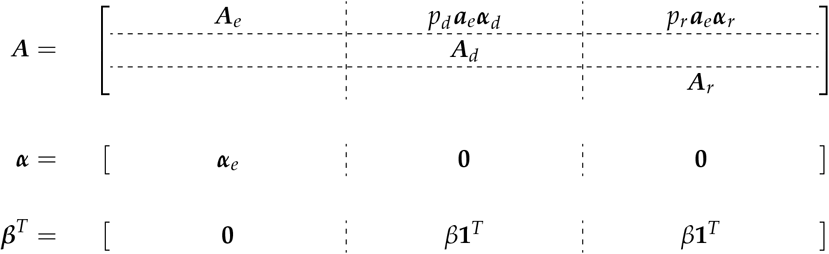

**Figure 3:**
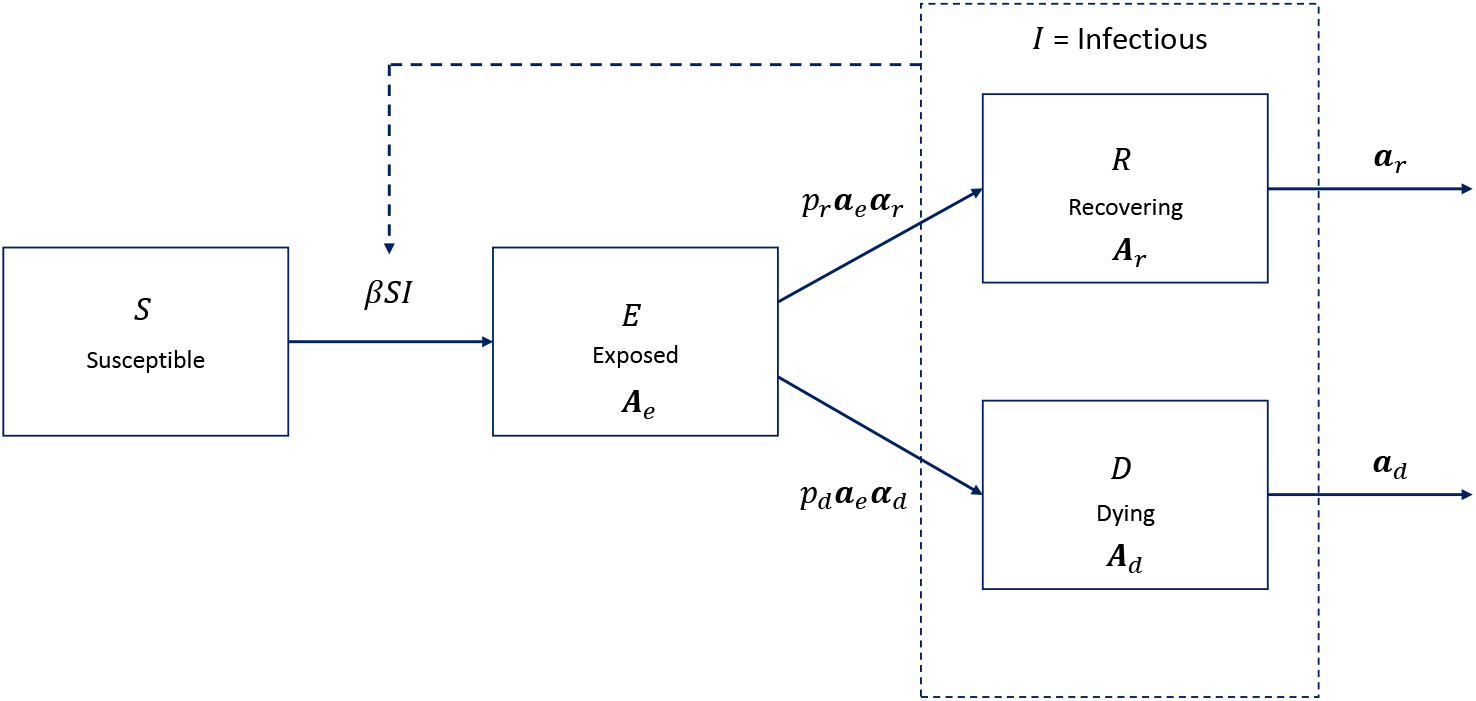
SEIRD Model.

The empty spaces denote matrix or vectors of zeros, of the appropriate size, and **1** are column vectors of ones, and ***β**^T^* is the transpose of column vector ***β***.

Figure 4 shows a numerical example of this model. The continuous lines show the result using the classical model, whereas the dotted lines show the model using Erlang distributions. We can see that the number of deaths and recovered in both models is the same, but the peak of infected individuals is higher with the Erlang distributions are used.

**Figure 4:**
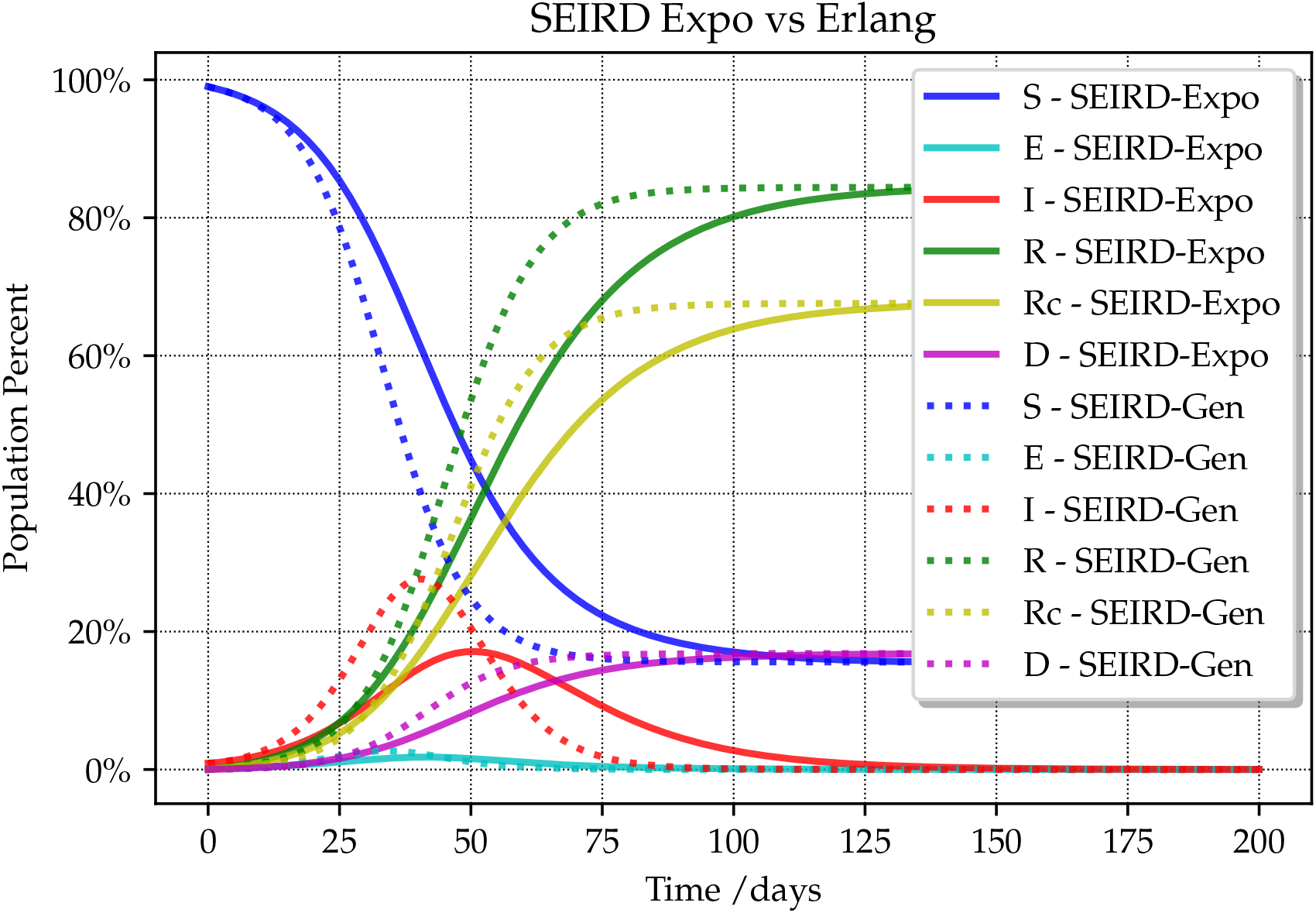
SEIRD Model: comparisoon of results with exponential and with Erlang distributions.

## 7 Limit Behavior of SIR-G Models

In the classical SIR model, the percentage of individuals that ever get infected can be obtained by finding the value *R*_∞_ that solves the following equation [3]

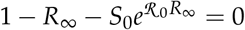

This value is very important for public policy, because it can be used to estimate the number of people that would eventually die.

We will now show that the same equation applies for the SIR-G, SIR-PH and SIR-PHG, by redefining *R*_0_ as explained below. We will prove the result for the SIR-PHG model; the SIR-PH will be a particular case and, since any distribution can be approximated with a PH, this will prove that the result also holds for the SIR-G model.

From (21c), we have

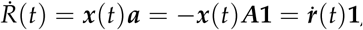

where ***ṙ***(*t*) *≡ −**x***(*t*)***A***. Since ***A*** always has an inverse, then ***x***(*t*) = *−**ṙ***(*t*)***A**^−^*^1^. Plugging this into (21a), we get

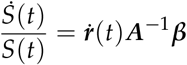

We can think of *S* as a function of ***r***, so integrating both sides we get

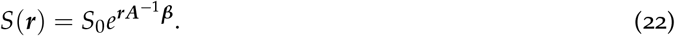

In the limit, *S*(***r***) *→* 1 *− R*_∞_, since there will be no infectious individuals left, so

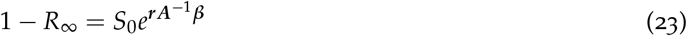

To analyze the behavior of ***r*** in the limit, write the vector ***x***(*t*) as

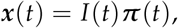

where the *i*-th component of ***π***(*t*) is the fraction of infectious individuals currently in phase *i*. In the limit, that vector goes to a vector ***π*** that is the steady-state solution of a CTMC with generator given by ***Q*** = (***A*** + ***αa***). This process is built assuming that, upon absorption from state *i* (with rate *a_i_*), a new phase *j* is immediately selected (with probability *α_j_*). See [12]. In other words, ***π*** is the solution to

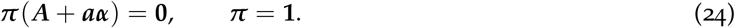

If you build a renewal process with time between arrivals *PH*(***a, A***), then the long term rate of arrivals is *γ* = 1/ E [*T*]. It can also be computed as *γ* = ***πa***, since it can be seen as long-term reward in a Markov process with reward vector ***a***. Using (24), we can establish these two equations that will be used in the sequel

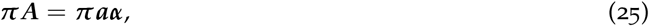

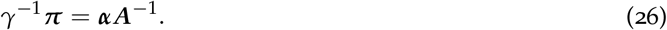

Using this machinery, in the limit we have

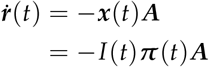

Now, *Ṙ*(*t*) = ***ṙ***(*t*)**1** = *−I*(*t*)***π***(*t*)***A1*** = *I*(*t*)***π***(*t*)***a***, so, using (25),

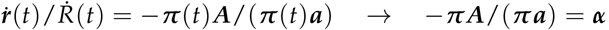

In the limit ***r***(*t*) will have the same direction as ***ṙ***(*t*), so

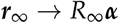

Plugging this into (22) we get

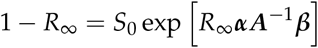

Finally, notice that from (26), ***αA***^−1^***β*** = *γ*^−^***πβ***. Putting all together, in (23) we get the desired result

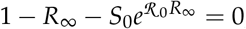

where

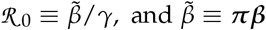

Here, 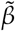 can be interpreted as a long-term weighted average infection rate, and *ℛ*_0_ will be an *effective* reproduction number for this system. Notice that in the SIR-PH model, these results reduce to ***β*** = ***β1***, and *R*_0_ = *β*/*γ*, as in the classic SIR. This shows that SIR-G model will behave in the long run identically to the classical SIR.

## 8 Other Generalizations

So far in this paper, we presented generalizations of the SIR model. However, it should be realized that the techniques presented can be generalized to many other models like SIS, etc.

For example, consider a SIS model. In such a model, infectious individuals always recover and become susceptible again. Then an SIS-G model in integro-differential form would be

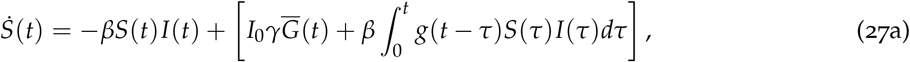

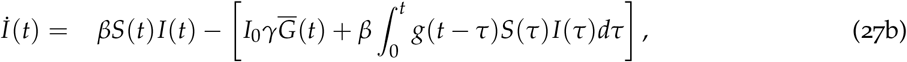

subject to *S*(0) = *S*_0_ and *I*(0) = *I*_0_. Since *I*(*t*) + *S*(*t*) = 1, this model can be converted to a single function, but it is not obvious that doing that you could obtain an analytical solution, as in the classical SIS. Using PH distributions, the equivalent model would be

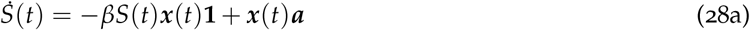

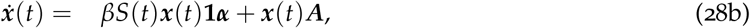

subject to ***x***(0) = *I*_0_***α***^′^ and *S*(0) = *S*_0_. Again, the model can be written only in terms of *bx*(*t*) only, since *S*(*t*) = 1 *− **x***(*t*)**1**.

We did not explore the numerical properties of these type of models.

## 9 Conclusions

In this paper, we have presented diverse epidemics models with arbitrary probability distributions for the infection period. We have shown the effect of different distributions in the dynamics of the epidemic.

Our focus has been in the SIR-like models, but the techniques can be applied to other models. Further research is needed to see the impact of variability in other types of models.

Finally, it remains to be seen how different results can be obtained by fitting data from a real-life pandemic, like the currently on-going COVID-19.

## Data Availability

Data and Code available at https://github.com/griano/epistoch

## A Appendix: Computing Expected Values of Piece-wise Linear Functions

In this appendix we will establish an efficient formula to compute the integral

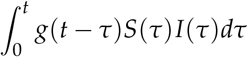

By a change of the integration variable (or just noticing that this is a convolution), it can be shown that this integral is equivalent to

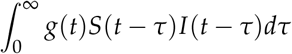

The limit of the integral can be changed, since we can make sure *S*(*t*)*I*(*t*) = 0 for *t* < 0. The previous integral can be seen as the expected value of a function, E [ *f* (*T*)] where, for a fixed *t*, we have *f* (*x*) = *I*(*t − x*)*S*(*t − x*).

Therefore, we will build a general formula to compute the expected value of a piece-wise linear function of a random variable in terms of the survival function, 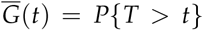, and the first order loss function *L*(*t*) = E [(*T − t*)^+^]. The advantage of this representation is that the loss function is known for many distributions, and can be efficiently computed without performing actual integration. See, e.g., [18, Page 14] and [19, Appendix C].

Split the time line in time 0 *≡ t*_0_ < *t*_1_ < …. Consider a piece-wise linear function given by

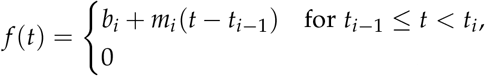

See Figure5. In what follows, we will use the following notation: For any array *b_i_* we define *∆b_i_* = *b_i_*_+1_ *− b_i_*, the indicator function 1(*A*) is 1 if statement *A* is true, and zero otherwise, *δ_i_* = *∆t_i_* = *t_i_*_+1_ *− t_i_*, for any real quantity *x*^+^ = max*{x*, 0*}*; note that (*t − a*)1(*t > a*) = (*t − a*)^+^. We will define *s_i_* to be size of the step at *δ_i_*, i.e., *s_i_ ≡ b_i_*_+1_ *−* (*b_i_* + *m_i_*(*t_i_ − t_i−_*_1_)) = *∆b_i_ − m_i_∆t_i_*. (See Figure5). Using this, we can write *f*(*·*) as

**Figure 5:**
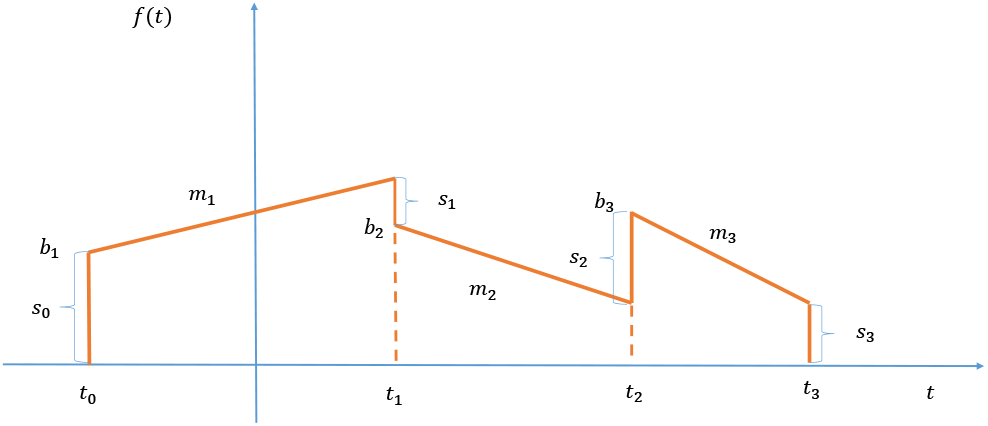
Piece-wise linear function

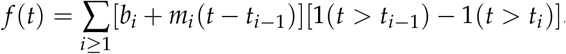

We now separate the two sums and rearrange terms

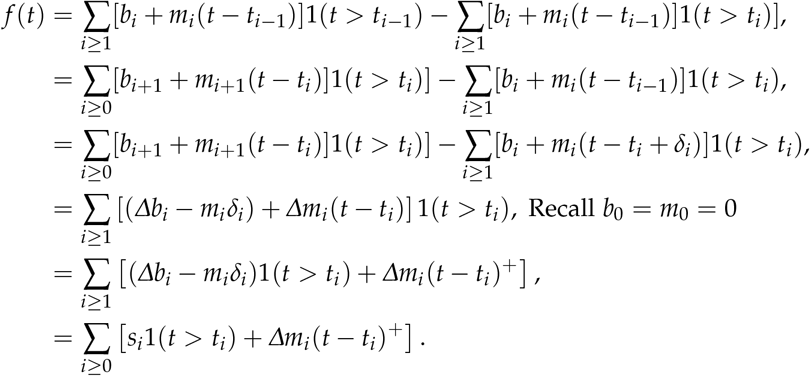

Taking expected value in the previous expression, and defining 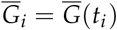 and *L_i_* = *L*(*t_i_*), then

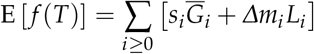

If the function is piece-wise continuous, then *s_i_* ≥ 0, except for *s*_0_ = *b*_1_, and the slopes are *m_i_* = *∆b_i_*/*δ_i−_*_1_. Hence, in this case,

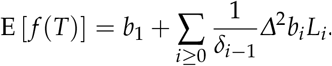

Notice that all the *G_i_* and *L_i_* can be pre-computed.

The integral that appears in (8)

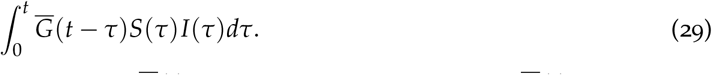

This integral can be computed in a similar fashion. 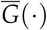 is not a density. However, 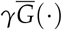 is a density, with 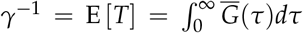. In fact, it is the density that corresponds to the residual equilibrium distribution in (4). Therefore, the integral (29) can be thought of as E [*f* (*T*^0^) with *T*^0^ having survival function given by

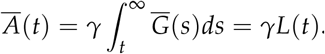

The loss function for *T*^0^ is

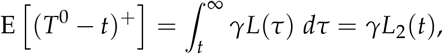

where *L*_2_(*t*) is the second order loss function of the original variable, namely,

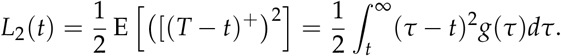

This function is readily available for many common distributions.

